# Neopterin level can be measured by intraocular liquid biopsy

**DOI:** 10.1101/2022.07.01.22276646

**Authors:** Anelia Kuvbachieva-Benarrosh, Chaker Nefzaoui, Isabelle Quadrio, Charles-Henry Remignon, Eric Jullian, Armand Perret-Liaudet

**Affiliations:** Ophthalmology Department, Sainte Musse Hospital, 54 avenue Henri Sainte-Claire Deville, 83100 Toulon, France; Neurochemistry and Neurogenetics Unit, Department of Biochemistry and Molecular Biology, Lyon University Hospital, 69677 BRON Cedex, France; Biochemistry Department, Sainte Musse Hospital, 54 avenue Henri Sainte-Claire Deville, 83100 Toulon, France

**Author notes:** Corresponding Author: Anelia Kuvbachieva-Benarrosh, Neurology Department, Sainte Musse Hospital, 54 avenue Henri Sainte-Claire Deville, 83100 Toulon, France.

## Abstract

Robust intraocular markers are needed in clinical practice for several inflammatory ophthalmological conditions in addition to advanced imaging, histology and immunohistochemistry tests in order to assure reliable diagnosis. Neopterin (NPt) is produced by activated lymphocyte T cells and is known to increase within the CNS in proinflammatory or early infectious states. This study aimed to measure intraocular NPt in aqueous liquid following intraocular liquid biopsy using standard methods. We enrolled 20 healthy patients without known inflammatory history or medication underdoing cataract surgery and analyzed samples for 19 out of 20. NPt level was measured below 1,9 nmol/l in 16 patients (84 %) and less than 3 nmol/l in 94,74%. Given the first time that NPt is measured in intraocular liquids we compared with the established reference limits within the CSF. These findings suggest that NPt could serve as a potential additional signature in ophthalmological conditions to differentiate between any or proinflammatory intraocular state, as well in follow-up.

## Main Text

Neopterin (NPt) is a pteridine derivative synthesized by macrophages and monocytes upon activation by cytokines, mostly interferon-gamma, revealing a proinflammatory immune condition. Npt is also produced by increased oxidative stress, following immune-response increased production of reactive oxygen species.

Within the central nervous system (CNS), high Npt levels were already reported in viral or bacterial meningoencephalitis as well some brain tumors or in immune-mediated neuroinflammatory conditions. It is largely accepted that the immune response within the CNS is guided by lymphocyte T helper cells type 1 (1). There is evidence that the CNS Npt is produced also by microglia and astrocytes, triggered by interferon-gamma. It is considered that plasma and CNS Npt levels are not correlated, suggesting brain immune activation is independent on the blood-CSF barrier dysfunction (2). Initially, CSF Npt was studied in Human Immunodeficient Virus (HIV) patients (3). During their clinical improvement, it was found a positive correlation between high Npt levels, severity of immunosuppression within the CNS and blood CD4 lowering. More generally, CSF Npt seems to be systematically elevated in patients exhibiting a viral encephalitis. To date, there is no report of absence of elevation of CSF Npt in acute or chronic viral etiology.

It has been determined for immunocompetent patients a cut-off value for the CSF neopterin allowing to differentiate CNS immune cellular activation and non-cellular immune conditions, acute or chronic. However, neopterin remains a helpful marker in primary CNS lymphoma diagnostic strategy (4).

Liquid biopsy in ophthalmology is performed in both aqueous and vitreous humor for biochemical and/or cell count for diagnostic or treatment options.

Our study aimed to detect and quantify intraocular NPt in aqueous liquid of patients with anterior chamber surgery, as the cataract. The procedure followed national recommendations and established protocol (clear corneal micro-incisions, intracameral dispersive viscoelastic administration, capsulorhexis, hydrodissection, emulsification of the nucleus, cortical cleanup, cohesive viscoelastic injection in the capsular bag, intraocular lens implantation, hydrosuture of incisions, intracameral antibiotic injection). The surgery is usually performed under topical or local anesthesia and the patient can be discharged the same day. Postoperative follow up is usually on day 1, week 1, and week 4.

In order to exclude false positive levels of intraocular Npt and because of lack of reference values, we performed urine test. We adjusted urine neopterin/creatinine ratio upon the reference values of the age and sex.

We included 20 immunocompetent patients admitted in our hospital for standard cataract surgery, aged from 55 to 89, mean age 75,55 (SD 8,2), 10% less than 65 years, equal sex ratio.

No one had glaucoma, infection, systemic vasculitis or granulomatosis. Three patients had diabetes. One had renal failure.

We analyzed 19 out of 20 patients with a possibility to check, if necessary, on a second aliquot. After collection in vials to protect Npt against UV oxidation, aqueous liquid was frozen at −80°C before analysis.

Mean volume of the aqueous liquid was 264 (SD 80,94).

Measurement of Npt concentrations in aqueous liquid and urine was performed using high-performance liquid chromatography coupled with fluorimetric detection (Waters). Pools of CSF were used as internal quality control with an interassay coefficient of variation of 5.9% and 7 % for 7.5 and nmol/L levels respectively. For creatinine measurement in urine, an enzymatic determination was used (C16000 analyzers, Abbott). Npt was measured in 19 subjects. In 16 patients (84%) Npt level was below 1,9 nmol/l and in 94,74% it was less than 3 nmol/l. The only one measurement at 4,1 nmol/l was in a subject with renal failure, but we could not perform a second analysis because of lack of enough amount of aqueous liquid. Intraocular neopterin levels less than 3 nmol/l could be compared to those found in CSF where we have very well-known established reference limits (5).

In conclusion, our study shows that intraocular Npt can be measured in adult patients using liquid biopsy even if we do not have statistically significant cut-off level. Furthermore, we consider that a volume less than 100 µl would be a minimum volume for a study in order to respect methodological limits. Additionally, Npt is a robust and could support very low temperatures (storage at −80°C). Looking forward for a new intraocular biomarker, Npt could be used in inflammatory conditions where a significant correlation among the biomarker levels and the disease severity.

We would perform another study on a larger scale of subjects in order to be able to determine the dosage of intraocular Npt with a negative predictive value and corresponding sensitivity and specificity.

## Data Availability

Medical anonymized data for each patient enrolled in our study and relevant synthesis and statistical analysis.

## Authors’ Disclosures or Potential Conflicts of Interest

No authors declared any potential conflicts of interest.

## Notes

### Competing Interest Statement

The authors have declared no competing interest.

### Clinical Trial

NCT03497481

### Funding Statement

Centre Hospitalier Intercommunal Toulon La Seyne, 54 avenue Henri Sainte-Claire Deville, 83100 Toulon, Var, France

### Author Declarations

COMITE de PROTECTION DES PERSONNES Ile de France XI approved our work on September 21, 2018 on the behalf the ethics oversight

